# Expanding cancer predisposition genes with ultra-rare cancer-exclusive human variations

**DOI:** 10.1101/2020.01.09.19015867

**Authors:** Roni Rasnic, Nathan Linial, Michal Linial

## Abstract

It is estimated that up to 10% of cancer incidents are attributed to inherited genetic alterations. Despite extensive research, there are still gaps in our understanding of genetic predisposition to cancer. It was theorized that ultra-rare variants partially account for the missing heritable component. We harness the UK BioBank dataset of ∼500,000 individuals, 14% of which were diagnosed with cancer, to detect ultra-rare, possibly high-penetrance cancer predisposition variants. We report on 115 cancer-exclusive ultra-rare variations (CUVs) and nominate 26 variants with additional independent evidence as cancer predisposition variants. We conclude that population cohorts are valuable source for expanding the collection of novel cancer predisposition genes.

## Introduction

Discovery of cancer predisposition genes (CPGs) has the potential to impact personalized diagnosis and advance genetic consulting. Genetic analysis of family members with high occurrences of cancer has led to the identification of variants that increase the risk of developing cancer [1]. In addition to family-based studies, efforts to identify CPGs focus on pediatric patients where the contribution of environmental factors is expected to be small. Forty percent of pediatric cancer patients belong to families with a history of cancer [2].

Tumorigenesis results from mis-regulation of one or more of the major cancer hallmarks [3]. Therefore, it is anticipated that CPGs overlap with genes that are often mutated in cancerous tissues. Indeed, CPGs most prevalent in children (TP53, APC, BRCA2, NF1, PMS2, RB1, and RUNX1) are known cancer driver genes that function as tumor suppressors, oncogenes or have a role in maintaining DNA stability [4]. Many of the CPGs mutated in somatic tissues are associated with DNA-repair pathways and homologous recombination [5]. The inherited defects in cells’ ability to repair and cope with DNA damage are considered as major factors in predisposition to breast and colorectal cancers [6].

Complementary approaches for seeking CPGs are large-scale genome / exome wide association studies (GWAS) which are conducted solely based on statistical considerations without prior knowledge on cancer promoting genes [7]. Identifying CPGs from GWAS is a challenge for the following reasons: (i) Limited contribution of genetic heritability in certain cancer types; (ii) Low effect size / risk associated with each individual variant; (iii) Low-penetrance in view of individual’s background [8], and (iv) Low statistical power. Large cohorts of breast cancer show that ∼2% of cancer cases are associated with mutations in BRCA1 and BRCA2 which are also high-risk ovarian cancer susceptibility genes. Additionally, TP53 and PTEN are associated with early-onset and high-risk familial breast cancer. Mutations in ATM and HRAS1 mildly increase the risk for breast cancer but strongly increase the risk for other cancer types and a collection of DNA mismatch repair genes (MLH1, MSH2, MSH6, PMS2) are associated with high risk of developing cancer [9]. A large cohort of Caucasian patients with pancreatic cancer reveal 6 high risk CPGs that overlap with other cancer types (CDKN2A, TP53, MLH1, BRCA2, ATM and BRCA1) [10].

Estimates for the heritable component of predisposition to cancer were extracted from GWAS, family-based and twin studies [11-13]. These estimates vary greatly with maximal genetic contribution associated with thyroid and endocrine gland cancers, and a minimal one with stomach cancer and leukemia [14]. Current estimates suggest that as many as 10% of cancer incidents can be attributed to inherited genetic alterations (e.g., single variants and structural variations) [15, 16]. The actual contribution of CPGs varies according to gender, age of onset, cancer types and ethnicity [17-20]. It is evident that high risk variants with large effect sizes are very rare [21]. Actually, based on the heritability as reflected in GWAS catalog, it was estimated that only a fraction of existing CPGs is presently known [22]. Therefore, instances of extremely rare mutations with high risk for developing cancer remain to be discovered.

A catalog of 114 CPGs was compiled from 30 years of research [1] with about half of the reported genes derived from family studies representing high-penetrance variants. An extended catalog was reported with a total of 152 CPGs that were tested against rare variants from TCGA germline data, covering 10,389 cancer patients from 33 cancer types and included known pediatric CPGs [23]. The contribution of BRCA1/2, ATM, TP53 and PALB2 to cancer predisposition was confirmed.

In this study we report on known and novel cancer predisposition candidate genes. We benefit from the UK-Biobank (UKBB), an invaluable resource of germline genotyping data for ∼500,000 individuals. The UKBB reports on ∼70,000 cancer patients and ∼430,000 cancer free individuals, considered as control group. We challenge the possibility that CPGs can be identified from very rare events, henceforth called cancer-exclusive ultra-rare variants (CUVs). These CUVs are expected to exhibit high penetrance. Here we report on 115 exome variations, 72 of which are heterologous. The majority of the matching genes are novel CPG candidates. We provide indirect genomic support for some of the CUVs that occur within coding genes and discuss their contribution to tumorigenesis.

## Results

The primary UK Biobank (UKBB) data set used in the article is comprised of 325,407 Caucasian filtered UKBB participants (see Methods), 282,435 cancer-free (86.8%) and 42,972 diagnosed with at least one malignant neoplasm. Among participants with cancer, 55% were diagnosed with either skin or breast cancer. The ICD-10 codes assembly is summarized in Supplementary Table S1. A total of 13.2% of the cancer-diagnosed individuals had two or more distinct neoplasms diagnosed. The validation UKBB data set includes 70,544 non-Caucasian, filtered participants, 63,585 cancer free (90.1%). Fig. 1a and Fig. 1b provide further details on different cancer type prevalence in these sets.

**Figure 1.**
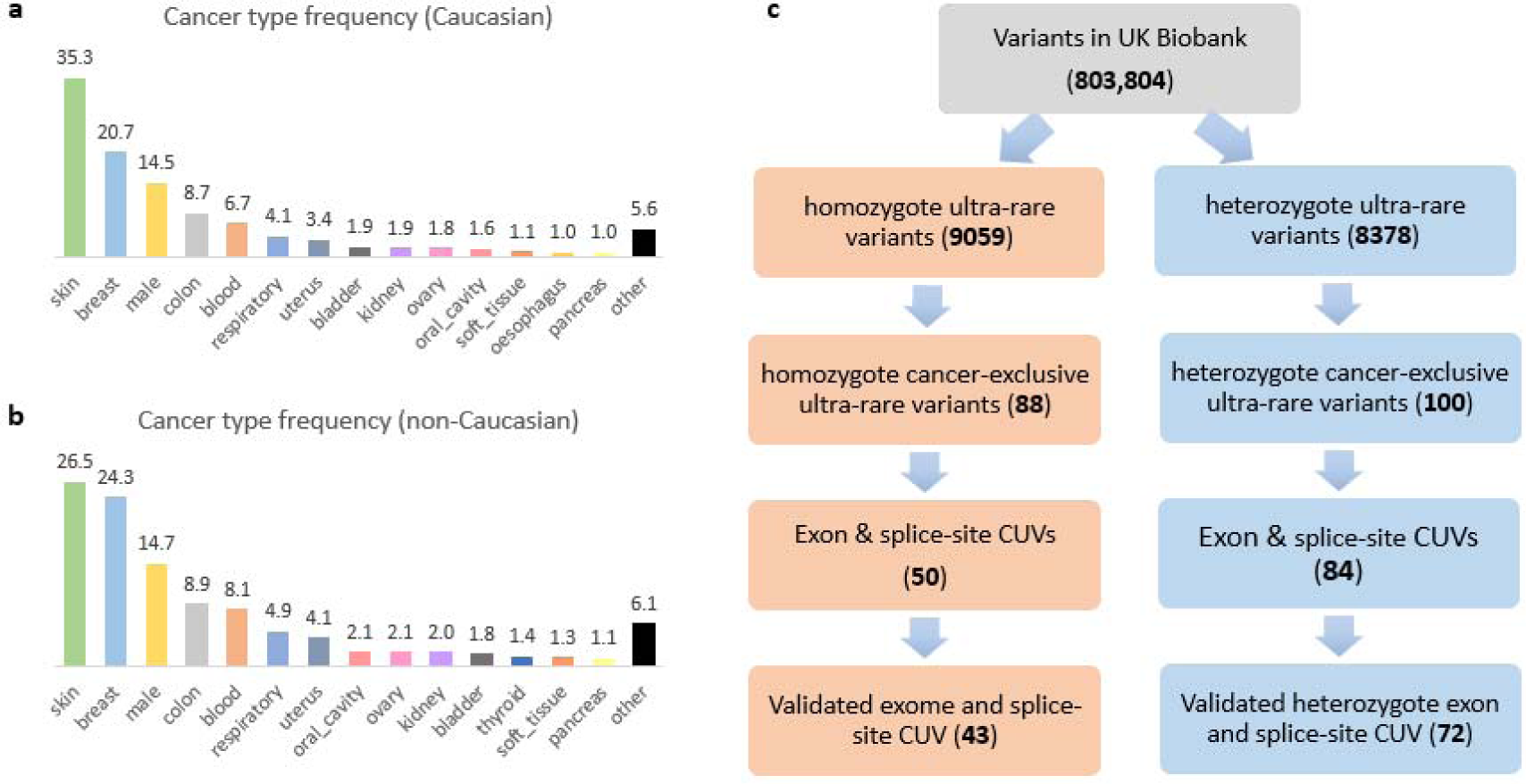
UK Biobank CUVs collection. The Caucasian filtered UK Biobank (UKBB) data set include 42,972 individuals who had cancer and the non-Caucasian filtered include 6,959 such individuals. **(a)** Cancer type distribution for the Caucasian data set. **(b)** Cancer type distribution for the non-Caucasian data set. **(c)** Out of 803,304 UKBB variants, we curated 72 heterozygous and 43 homozygous CUVs (total 115 CUVs).

### Compilation of cancer-exclusive ultra-rare variants (CUVs)

We scanned 803,804 genetic markers in our prime data set for cancer-exclusive variations. 188 variations met our initial criteria, appearing at least twice in individuals diagnosed with cancer and not appearing in cancer-free individuals. One hundred of the variations were heterozygous and 88 were homozygous. In order to target variations with additional supporting evidence, we considered only coding exome and splice-region variants. An additional rarity filtration performed using the gnomAD data set reduced the list to 115 variants (associated with 108 genes), 72 heterozygous and 43 homozygous (Fig. 1b). The detailed list of all 115 CUVs can be found in Supplementary Table S2.

Most (66%) of the CUV are missense variants. There is a strong enrichment for loss of function (LoF) variants (i.e., frameshift, splicing disruption and stop gains), which account for 33% of the CUVs. Only a single homozygous CUV is synonymous (Fig. 2a). The distribution of variation types varies greatly between homozygous and heterozygous CUVs (Fig. 2b). Missense variants are 93% of the homozygous variants, but only 50% of the heterozygous CUVs. The heterozygous CUVs are highly enriched for LoF variants which constitute the other 50%.

**Figure 2.**
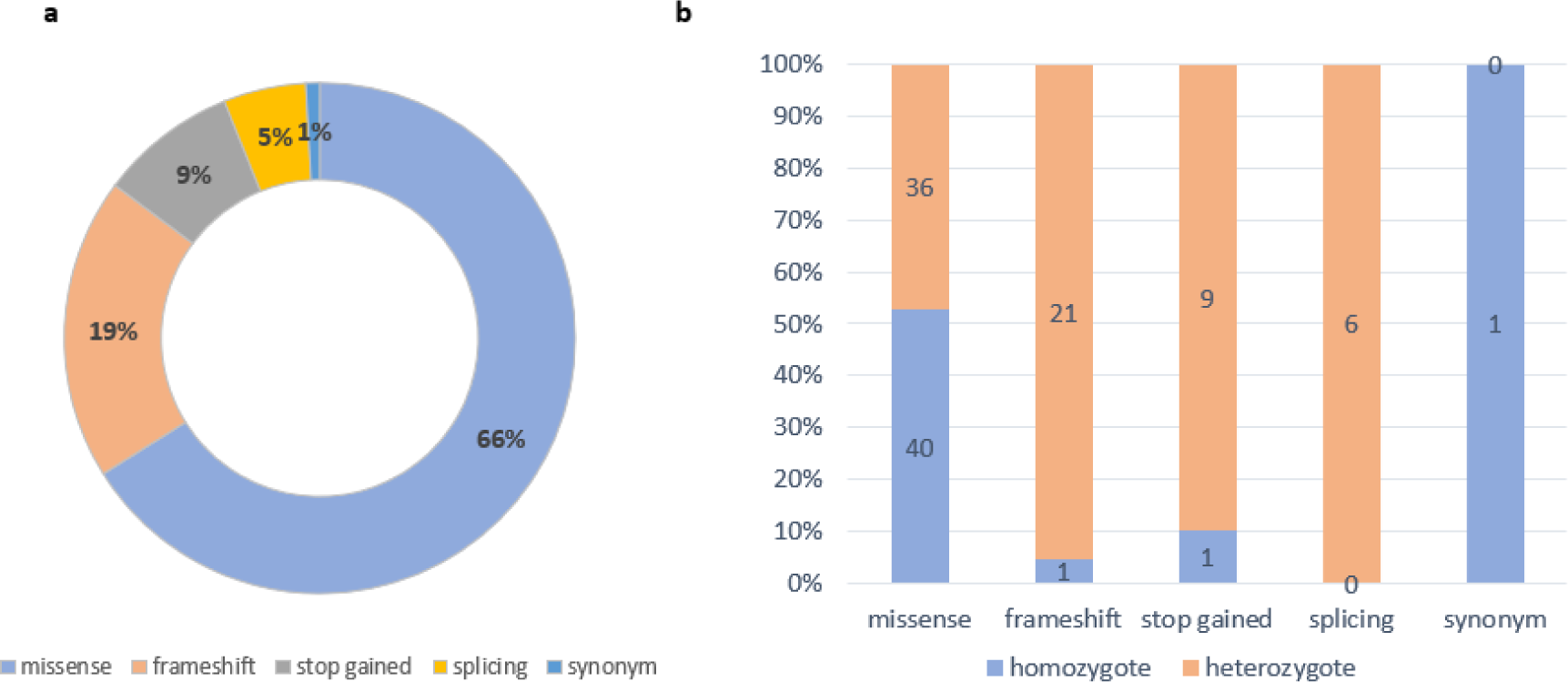
Exomic CUVs are mostly gnomically disruptive. The partition of variant types for the compiled list of 115 exomic CUVs. The list is dominated by transcript disruptive variations (99.1%) that include missense, frameshift, stop gain and splicing sites. **(a)** Distribution of variation types among the exomic CUVs. **(b)** Dispersion of variant type among heterozygous and homozygous CUVs.

### Cancer-exclusive ultra-rare variants (CUVs) overlap with known cancer predisposition genes

From the listed CUVs, 26 variants in 23 genes were previously defined as cancer inducing (Table 1). Specifically, 22 CUVs within 19 genes appear in the updated list of CPG catalog [23] and 24 CUVs within 21 genes are known cancer driver genes (Fig. 3a), as defined by either COSMIC [27] or the consensus gene catalog of driver genes (listing 299 genes, coined C299) [28]. More than half of the cancer associated variants result in loss of function (LoF). Many of the affected genes are tumor suppressor genes (TSGs), among which are prominent TSGs such as APC, BRCA1 and BRCA2 (Table 1), each identified by two distinct CUVs.

**Table 1.** CUVs overlap with known cancer predisposition or driver genes

| hg19 | Effect | Gene | COSMIC | C299 | CPG | Function<sup>a</sup> |
| --- | --- | --- | --- | --- | --- | --- |
| 1:155205517 | missense | **GBA** |  |  | Y | Enzyme |
| 2:48027130 | missense | **MSH6** | Y | Y | Y | DNA repair |
| 3:10183771 | missense | **VHL** | Y | Y | Y | Ubq-complex |
| 3:30730003 | splice region | **TGFBR2** | Y | Y |  | Kinase |
| 3:37048480 | splice region | **MLH1** | Y | Y | Y | TSG |
| 5:112173671 | frameshift | **APC** | Y | Y | Y | TSG |
| 5:112175255 | frameshift | **APC** | Y | Y | Y | TSG |
| 9:101891277 | stop gain | **TGFBR1** |  |  | Y | Kinase |
| 9:131341997 | missense | **SPTAN1** |  | Y |  | Cytoskeletal |
| 10:43609079 | frameshift | **RET** | Y | Y | Y | Kinase |
| 10:88659605 | missense | **BMPR1A** | Y |  | Y | Kinase |
| 10:89717630 | stop gain | **PTEN** | Y | Y | Y | TSG, Phosphatase |
| 11:44193237 | missense | **EXT2** | Y |  | Y | TSG, Enzyme |
| 11:71720337 | missense | **NUMA1** | Y |  |  | MT Spindle pole |
| 11:108192066 | missense | **ATM** | Y | Y | Y | DDR, Kinase |
| 13:32890621 | frameshift | **BRCA2** | Y | Y | Y | TSG, DNA repair |
| 13:32914296 | missense | **BRCA2** | Y | Y | Y | TSG, DNA repair |
| 13:48878061 | frameshift | **RB1** | Y | Y | Y | TSG |
| 13:103524611 | frameshift | **ERCC5** | Y |  | Y | DNA repair |
| 16:2121553 | missense | **TSC2** | Y | Y | Y | TSG |
| 17:29654601 | missense | **NF1** | Y | Y | Y | RAS regulator |
| 17:41244383 | frameshift | **BRCA1** | Y | Y | Y | TSG, DNA repair |
| 17:41246296 | stop gain | **BRCA1** | Y | Y | Y | TSG, DNA repair |
| 18:3451996 | frameshift | **TGIF1** |  | Y |  | TGF ligand |
| 21:36421256 | splice region | **RUNX1** | Y | Y | Y | TF |
| 22:30067894 | missense | **NF2** | Y | Y | Y | Cytoskeletal |

<sup>a</sup>Function abbreviation: DDR, DNA damage response; TSG, tumor suppressor gene; TF, transcription factor; MT, microtubule; Ubq, ubiquitin.

**Figure 3.**
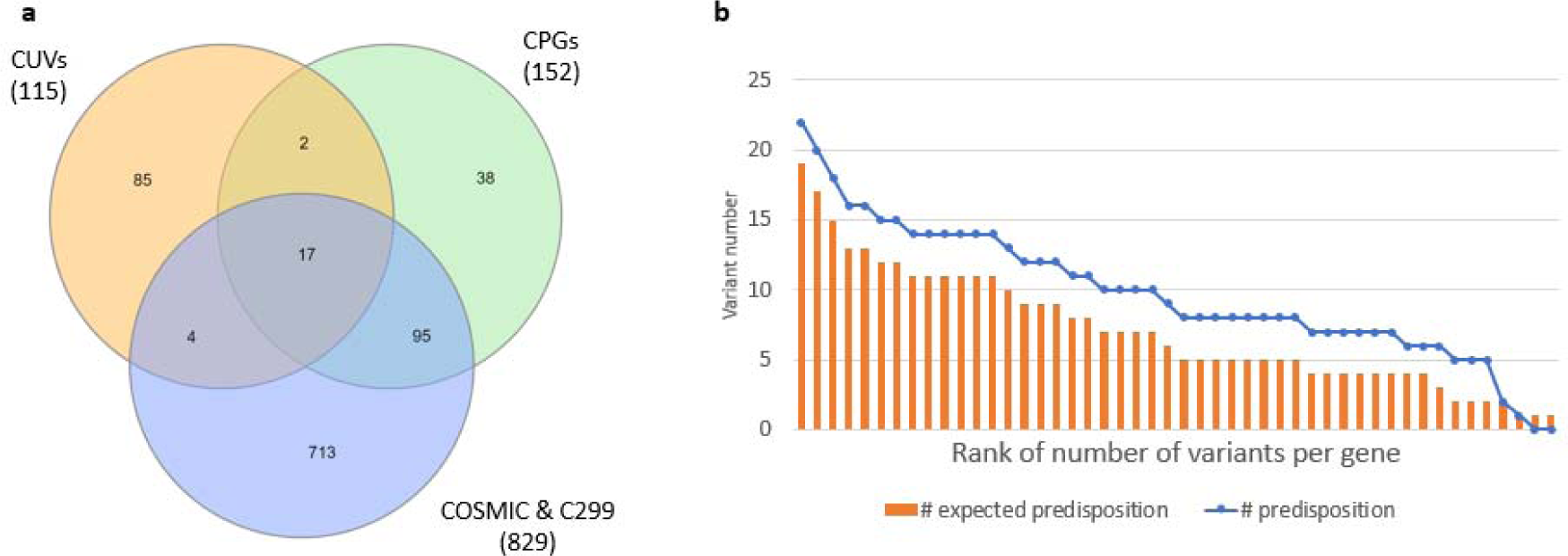
CUVs list is enriched with cancer predisposition genes. Out of the 108 genes in the CUVs list, 23 are known cancer genes. **(a)** Venn diagram of the genes associated with CUVs, known cancer driver genes (as reported in COMICS) and the consensus CPGs. **(b)** Expected number of known CPG CUV (orange) versus the actual number of known CPG in heterozygote CUVs (blue). An unbalanced representation of genes in ultra-rare variants of UKBB results in over-representation of some genes. We therefore ranked the genes based on number of ultra-rare variants (Supplementary Table S3). For each rank, we present the expected number of CUVs from CPGs and the actual number observed for CUVs from CPGs.

The heterozygous CUVs are enriched for known cancer predisposition genes. Twenty-five of the cancer associated CUVs are heterologous and one is homologous. Notably, there is an inherent imbalance in the initial variant sampling performed by the UKBB. As the UKBB use DNA arrays for obtaining genomic data, the identifiability of ultra-rare exome variants is restricted by the selection of SNP markers and the design of the array. There are 6450 heterozygous ultra-rare exome variants from 2938 genes which pass our biobank-ethnic and the gnomAD allele frequency filtration. A total of 1604 of the filtered ultra-rare variants are from 105 known CPGs, with some genes that are over-represented in the ultra-rare variants (Supplemental Table S3). For example, the exomic region of BRCA2 is covered by 226 such SNP marker variants, while most genes have none.

In order to account for the disproportional number of the ultra-rare variant of some CPGs, we calculated the expected number of cancer predisposed genes when gradually removing highly-represented genes from the ultra-rare variant collection for heterozygotes. As shown in Fig. 3b, there is an enrichment towards CPGs and even more so as we remove variants of over-represented genes (e.g., BRCA2). We calculated p-values for each data-point using a two-side binomial test. The results are available in Supplemental Table S3.

### Independent genetic validation

Due to the extremely rare nature of the CUVs, we require additional support for the collection of the CPG candidates. We seek independent genetic validation of the non-cancer related CUVs. We apply three sources for validation: (i) the filtered Caucasian UKBB cohort; (ii) the matched filtered, non-Caucasian UKBB cohort; (iii) the collection of germline variants from TCGA, as reported in gnomAD. The complete list of genetically validated novel CPG candidates is listed in Table 2.

**Table 2.** Novel validated CPG candidates

| Gene Symbol | zygote form | # people per CUV | Distinct CUVs | non-Caucasian cohort | TCGA germline | function in tumorigenesis | Ref. |
| --- | --- | --- | --- | --- | --- | --- | --- |
| AGR2 | hetero | 3 |  |  |  | Affects cell migration, transformation and metastasis. Wnt signaling, tumor antigen. | [34] |
| AKR1C2 | hetero | 2 |  |  | Y | Exerts an inhibitory effect on oncogenesis | [35] |
| DNAH3 | homo | 3 |  |  | Y | Cancer predisposed genes in Tunisian family | [36] |
| DSP | hetero | 2 | Y | | | Affects cell adhesion. Suppressed by TGF-β | |
| EGFLAM | hetero | 2 |  |  | Y | Promotes matrix assembly |  |
| ENDOU | homo | 3 |  |  |  | Cancer biomarker | [37] |
| HIST1H2BO | hetero | 3 |  |  |  | Affects major signaling pathways |  |
| HSPB2 | hetero | 2 |  |  | Y | Epigenetically regulated | [38] |
| ICAM1 | homo | 4 |  |  |  | Biomarker, under a clinical trial | [39] |
| ISLR | homo | 2 |  | Y |  | Marker for mesenchymal stem cells. Deregulated gene in cancer | [40] |
| KCNH2 | hetero | 2 | Y |  |  | Affects proliferation and migration |  |
| MAP3K15 | hetero | 2 |  |  | Y | Contributes to cell migration |  |
| MRPL39 | hetero | 2 |  |  | Y | Tumor suppressor by targeting miR-130 | [41] |
| MYBPC3 | both | 2 | Y |  |  | Cytoskeletal modifier |  |
| MYO1E | homo | 2 |  | Y |  | Stimulates upregulation of motility and invasion | [42] |
| NAV3 | hetero | 3 |  |  |  | Acts as a suppressor of breast cancer | [43] |
| PCDHB16 | homo | 3 |  | Y | Y |  |  |
| SARDH | homo | 2 |  | Y |  | Acts as tumor suppressor | [44] |
| SCN5A | hetero | 2 | Y |  |  | Promotes breast cancer, possess anti-pancreatic cancer | [45] |
| WDFY4 | hetero | 2 |  |  | Y | Presentats viral, tumor antigen on dendritic cells | [46] |
| ZFC3H1 | homo | 2 |  |  | Y | Indirect activating DNA repair |  |

Within the Caucasian cohort, we consider the following as additional genomic evidence: (i) a gene with 2 CUVs, or (ii) any CUV seen in more than two individuals diagnosed with cancer. We found 7 genes that have 2 distinct CUVs, 3 of which are already known CPGs: BRCA1, BRCA2 and APC. The other 4 genes are likely novel CPG candidates: DSP, KCNH2, MYBPC3 and SCN5A. There are 9 CUVs which we detected in three individuals with cancer. Three of them are known predisposition or driver genes: NF1, ATM and TGFBR2. The other 6 genes are CPG candidates that were not previously assigned as such. This set includes PCDHB16, DNAH3, ENDOU, AGR2, HIST1H2BO and NAV3. Interestingly, a certain homozygous CUV in the gene ICAM1 appeared in 4 individuals with cancer in our filtered Caucasian cohort.

The non-Caucasian UKBB cohort provides additional independent genomic evidence. There are 5 CUVs that appear at least once in an individual with cancer from the non-Caucasian cohort. CUVs from the genes MYO1E, SARDH and ISLR appeared in two distinct individuals with cancer from this non-Caucasian cohort, while CUVs from PCDHB16 and known CPG BMPR1A appeared in a single individual with cancer.

TCGA germline variants were obtained using exome sequencing and thus offer an additional separate source for CUV validation. Clearly, the appearance of CUVs in TCGA germline data is not anticipated, as we discuss variants that are ultra-rare in both UKBB and gnomAD. The TCGA collection within gnomAD includes only 7,269 samples. We identified 10 CUVs that were also observed in TCGA gnomAD germline data, one of a known cancer driver gene TGIF1, and 9 novel CPG candidates: PCDHB16, EGFLAM, AKR1C2, MAP3K15, MRPL39, DNAH3, WDFY4, HSPB2 and ZFC3H1.

Based on the above support, we compiled a list of validated CPGs which includes 21 genes that are novel CPGs. Among these genes 12 CUVs are heterozygous, 8 are homozygous and MYBPC3 has both heterozygous and homozygous CUVs. Two of these genes have multiple validation evidence. DNAH3 with a homozygous CUV which appears in 3 individuals with cancer in the Caucasian cohort and within TCGA germline variant collection. PCDHB16 with a homozygous CUV which appeared in 3 individuals in the Caucasian cohort, one individual in the non-Caucasian cohort and in the TCGA gnomAD resource. In addition, non-CPG cancer-driver genes with validated CUVs include TGFBR2 and TGIF1 that are also very likely CPG candidates.

Some of the prominent genes in our list were signified by additional independent studies. For example, a novel oncolytic agent targeting ICAM1 against bladder cancer is now in phase 1 of a clinical trial [38]. Additionally, DNAH3 was identified as novel predisposition gene using exome sequencing in a Tunisian family with multiple non-BRCA breast cancer instances [35].

### Somatic mutations in novel CPGs significantly decrease survival rate

There is substantial overlap between CPGs and known cancer driver genes (Fig. 3a). This overlap suggests that somatic mutations in validated CPG candidates may have an impact on patients’ survival rate. We tested this hypothesis for the 21 novel CPG candidates (Table 2) using a curated set of 32 non-redundant TCGA studies (compiled in cBioPortal [1]) that cover 10,953 patients. By testing the impact of alteration in the 21 novel CPGs in somatic data we expect to provide a functional link between the germline CPG findings and the matched mutated genes in somatic cancer samples. Altogether, 3,846 (35%) of the patients had somatic mutations in one or more of the genes. The median survival of patients with somatic mutations in these genes is 67.4 months, while the median for patients without somatic mutations in any of these genes is much longer (86.3 months). Applying the Kaplan-Meier survival estimate yields a p-value of 1.78e-4 in the Logrank test (Fig. 4a). The Kaplan-Meier disease/progression-free estimate was also worse for patients with somatic mutations in the 21 novel CPGs with a p-value of 6.03e-3 (Fig. 4b).

**Figure 4.**
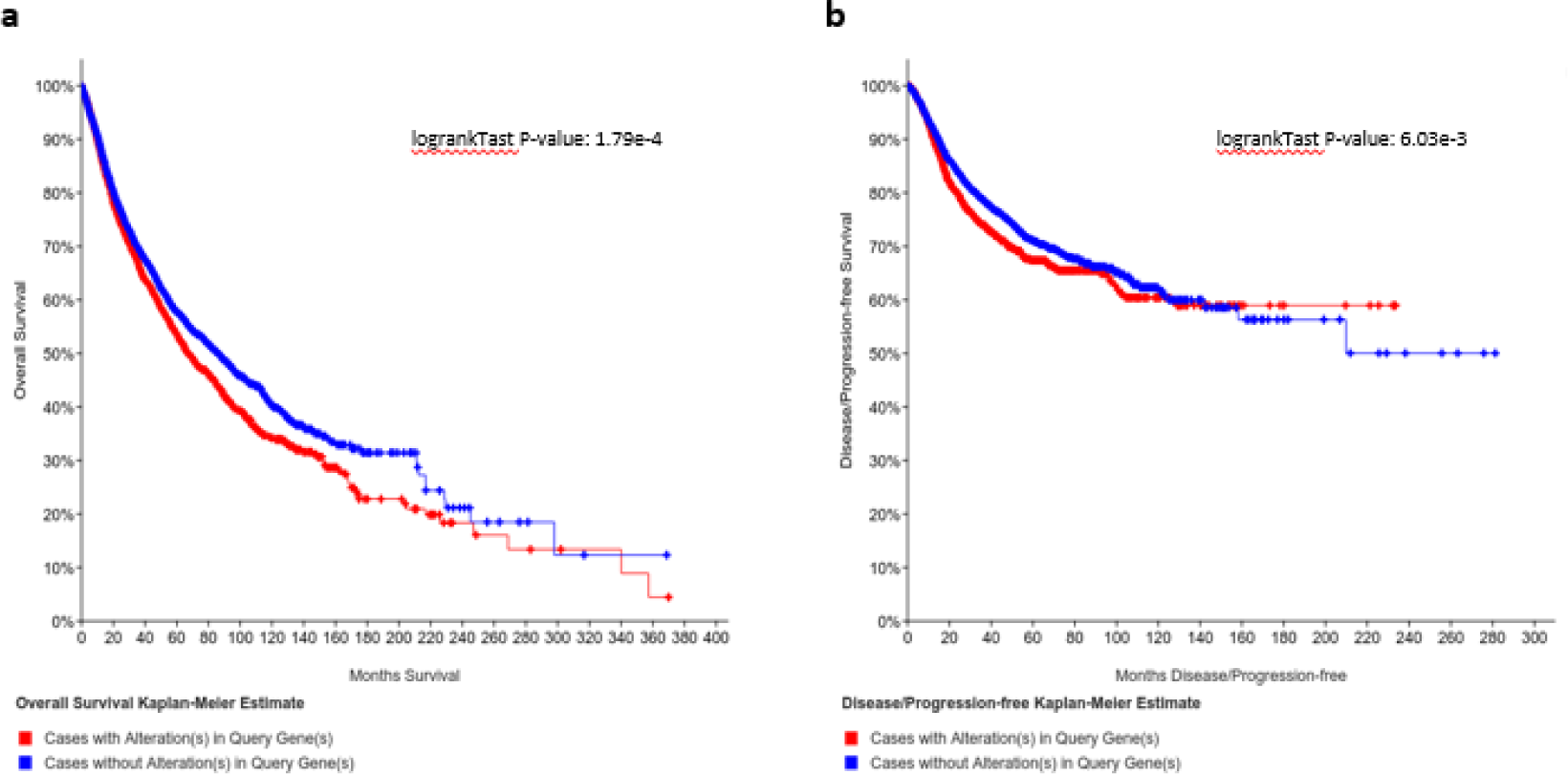
Somatic mutations in CPG candidate effect cancer patient survival and disease progression. The effect of somatic mutations in the 21 novel CPG candidate (Table 2) on the somatic mutations from TCGA cancer patients was tested (via cBioPortal). **(a)** Meier-Kaplan survival rate estimate. **(b)** Meier-Kaplan Disease / Progression-free estimate.

We conclude that the CUV-based CPG candidate genes from UKBB carry a strong signature that is manifested in patients’ survival, supporting the notion that these genes belong to an extended set of previously overlooked CPGs.

### Homozygous variations are mainly recessive

In order to ascertain whether the homozygous variations found are indicative of the heterozygous form of the variant as well, we viewed the heterozygous prevalence within the UKBB Caucasian population. In only a single variant in the gene MYO1E was the prevalence in healthy individuals significantly lower than in individuals with cancer (p-value: 0.04). As most of the variations have a strong cancer predisposition effect as homozygous variations, it seems that their influence is explained by a recessive inheritance mode. This phenomenon might explain the significant depletion of known CPGs within the homozygous variations in our list.

Inspecting the heritability model of previously reported CPGs [1] is in accord with our findings, showing that while about two-thirds of the genes comply with a dominant inheritance, the rest are likely to be recessive. Notably, in the most updated CPG catalog, 15% of the genes were assigned with both inheritance patterns. In our ultra-rare list, only MYBPC3 is associated with both heterozygous and homozygous variations.

## Discussion

We present a list of 115 CUVs from 108 genes. Among them 24 variants (from 21 genes) are associated with known cancer genes. Most of these variants (21) overlap with known cancer predisposition genes. Expanding the number of currently identified CPGs is crucial for better understanding of tumorigenesis and identifying various processes causing high cancer penetrance. Genetic consulting, family planning and appropriate treatment is a direct outcome of an accurate and exhaustive list of CPGs.

Known cancer predisposition variants only partially explain the cases of inherited cancer incidents. CPGs identification has already impacted cancer diagnostics, therapy and prognosis [1]. Genomic tests and gene panel for certain cancer predisposition markers are commonly used for early detection and in preventative medicine [29, 30]. It is likely that CPGs based on ultra-rare variants are not saturated. For example, additional CPGs including CDKN2A, NF1, and NBN were associated with an increased risk for breast cancer [31]. Specifically, CDKN2A has been also detected as a CPG in families of patients with pancreatic cancer [32].

With the impeding availability of UKBB exome sequencing (150,000 exomes), we will be able to revisit the identified variants and further refine the list of candidate CPGs (i.e., remove false-positives and add evidence to support true CPGs). Inspecting the function of genes associated with the 108 identified genes further supports the importance of protein modification (e.g. kinases and phosphatase function), chromatin epigenetic signatures [33], membrane signaling, DNA repair systems and more.

The inheritably rare nature of CUVs raise doubts on the reliability of their initial identification [26]. Therefore, we only considered as candidate CPGs those genes that are supported by additional independent genomic evidence from either the UKBB or the TCGA cohort. We nominate 23 genes as CPG candidates, two of which are known cancer drivers. As we have shown (Fig. 4), somatic mutations in the non-driver validated CPG candidates resulted in a significant negative effect on the patients’ survival rate.

## Materials and Methods

### Study population

The UK Biobank (UKBB) has recruited ∼500,000 people from the general population of the UK, using National Health Service patient registers, with no exclusion criteria [24]. Participants were between 40 and 69 years of age at the time of recruitment, between 2006 and 2010. To avoid biases due to familial relationships, we removed 75,853 samples keeping only one representative of each kinship group of related individuals. 312 additional samples had mismatching sex (between the self-reported and the genetics-derived) and 726 samples had only partial genotyping. We restricted our initial exploratory analysis to individuals that were genetically verified as Caucasians who also declared themselves as ‘white’. The rest of the participants not matching this criterion were used for the complementing analysis. The filtered Caucasian cohort includes 325,407 individuals (42,972 of whom had cancer) and the filtered non-Caucasian cohort includes 70,544 individuals (6,959 had cancer).

Additional data from gnomAD was used for variant rarity filtering and TCGA-germline validation [25].

### Rare variants reliability

Our CUV collection includes variants that appeared at least twice in the filtered Caucasian cohort, thereby evading many SNP-genotyping inaccuracies [26]. We further ascertain the validity of prominent variants with additional genomic evidence.

### Cancer type definition

The UKBB provides an ICD-10 code for each diagnosed condition. We considered an individual diagnosed with malignant neoplasm (ICD-10 codes C00-C97) as individuals with cancer, and otherwise as cancer-free individuals. The codes were aggregated to improve data readability using the assembly described in Supplementary Table S1.

#### Ethical approval

UK Biobank approval was obtained as part of the project 26664. Ethical approval for this study was obtained from the committee for ethics in research involving human subjects, for the faculty of medicine, The Hebrew University, Jerusalem, Israel (Approval number - 13082019).

## Data Availability

Most of the data that support the findings of this study are available from the UKBB. However, restrictions apply to the availability of these data, which were used under license for the current study, and so are not publicly available. Data are available from the authors upon a justified request and with permission of the UKBB. Data extracted from gnomAD is available from the authors upon request.

## Acknowledgement

We would also like to thank Nadav Brandes from the School of Computer Science and Engineering at the Hebrew University of Jerusalem for useful discussion and valuable comments. We thank Irene Unterman from the Medical School at the Hebrew University of Jerusalem for reading the manuscript. We thank the CSE system at the Hebrew University of Jerusalem team for their technical support.

## Competing Interests Statement

The authors declare that they have no competing interests.

## Supplementary files

**Table S1**. ICD10-code assembly

**Table S2**. The collection of 115 exome CUVs.

**Table S3**. Number of rare variants per gene, p-values for expected vs. observed CUVs from CPGs.

## Notes

### Competing Interest Statement

The authors have declared no competing interest.

### Funding Statement

No external funding was received for the presented work.

